# Childhood and adolescent cancer diagnosis, treatment, and hospitalizations during the COVID-19 pandemic in Québec, Canada, a population-based study

**DOI:** 10.64898/2026.11.18.26367724

**Authors:** Callum Mullen, Ronald D. Barr, Hallie Coltin, Erin Strumpf, Mariam El-Zein, Eduardo L. Franco, Talía Malagón

## Abstract

The COVID-19 pandemic disrupted healthcare delivery globally. In Québec, Canada, measures were introduced to prioritize pediatric oncology care. Using administrative data, we estimated pandemic-related changes in cancer diagnoses, treatments, and length of hospital stay among individuals aged 0–19 in Québec (2015–2023). Interrupted time series analyses using autoregressive integrated moving average (ARIMA) models assessed immediate and sustained changes in monthly age-standardized rates of diagnoses, treatments (chemotherapy, surgery, radiotherapy-related encounters), positron emission tomography scans, and hospital admissions. Separate ARIMA models fitted to pre-pandemic monthly counts generated counterfactual forecasts to estimate the cumulative differences between observed and expected diagnoses and treatments over 3, 12, and 46 months of the pandemic. Generalized linear mixed models assessed changes in length of stay between the pre- and post-pandemic periods and whether these changes differed by patient characteristics. We identified 2,570 patients: 1,475 diagnosed through February 2020 and 1,095 from March 2020 onward. We found no evidence of immediate or sustained changes in monthly incidence rates. Cumulative diagnoses and treatments remained within expected levels across periods, although estimates were imprecise. For diagnoses, the 3-, 12-, and 46-month cumulative differences were –1 (95% uncertainty interval [UI] –17 to 15), 24 (95% UI, –9 to 57), and 2 (95% UI, –94 to 96), respectively. Changes in length of hospital stay varied across subgroups, although their clinical significance remains uncertain. Pediatric cancer diagnosis and treatment activity was largely maintained across Québec throughout the pandemic, consistent with the objectives of provincial and institutional measures to preserve pediatric oncology services.

**Significance:** This population-based study provides a long-term assessment of pandemic-related changes in pediatric oncology care in Québec, Canada, extending earlier evidence focused on the initial pandemic period. The findings provide reassurance regarding the continuity of cancer diagnosis and treatment activity despite widespread healthcare disruptions.

## INTRODUCTION

Cancer remains a leading cause of disease-related death among children and adolescents worldwide.^1,2^ Although survival has improved substantially in recent decades, particularly in high-income settings, these gains depend on timely recognition of disease and access to specialized treatment.^3^ Disruptions in care pathways and healthcare-seeking behaviour, such as those introduced by the COVID-19 pandemic, raise concerns about the continuity of pediatric cancer care.

The pandemic’s immediate impacts were heterogeneous by region and far-reaching.^4^ Due in part to overburdened healthcare systems and risk mitigation strategies, reductions in cancer diagnoses among adult populations were documented across North America,^5–8^ Europe,^9,10^ and the United Kingdom,^11^ particularly for screen-detectable cancers.^5^ The impact of the pandemic on adult cancer diagnosis and care cannot be assumed to be equivalent for childhood cancer. Unlike adult cancers, population-based screening is not routinely implemented for childhood and adolescent cancers; diagnoses are driven by clinical presentation.^12,13^ Fewer healthcare encounters during the pandemic may have limited opportunities for signs and symptoms to be recognized and investigated. Notably, in the first year of the pandemic, family physician services among 0–17-year-olds decreased by 24% to 36% across four Canadian provinces.^14^ Even when care was sought, nonspecific symptoms may have been attributed to COVID-19 rather than to an underlying cancer, potentially delaying cancer diagnosis.^15^

Canadian evidence on the impact of the pandemic on childhood cancer care remains mixed. While no changes in the number of newly diagnosed childhood cancers were observed during the early pandemic period,^16^ analyses extending through 2022 reported fewer childhood cancers diagnosed overall,^17^ suggesting patterns may have differed as the pandemic evolved. Importantly, evidence regarding the delivery and continuity of cancer treatment among children and adolescents remains limited. Although a small survey of families receiving cancer care in Canada and the United States indicated that most treatments were unaffected, a minority reported changes to their type, timing, or delivery.^18^ A broader Canadian analysis also identified a 20% reduction in all cancer-related services during the first year of the pandemic, but did not provide pediatric-specific estimates.^19^ In Québec, governmental and institutional measures were implemented to protect pediatric oncology services.^20,21^ A central component of this response was preserving the specialized workforce delivering pediatric cancer care. Institutions were required to establish contingency plans for replacing absent medical and nursing staff with appropriately qualified personnel, while redeployment of oncology staff to acute-care settings was discouraged to limit transmission.^21^ The delivery of care during this period has not been well characterized at the population level in Québec. It therefore remains unclear whether childhood cancer diagnosis, treatment delivery, hospital admission, and length of stay changed during the pandemic, and whether any changes persisted beyond the early pandemic period.

Our objective was to estimate changes in the rates of childhood and adolescent cancer diagnosis and treatment delivery in Québec, Canada, attributable to the COVID-19 pandemic, and to assess differences in length of hospital stay before and during the pandemic.

## MATERIALS AND METHODS

### Study design and data sources

We conducted a retrospective, population-based cohort study using administrative health data from Québec, Canada, a province where a publicly funded healthcare system covers over 95% of the population and delivers medically necessary physician and hospital services.^22^ We linked records from the *Maintenance et exploitation des données pour l’étude de la clientèle hospitalière* (MED-ECHO) database, which captures all acute-care hospitalizations and day surgeries, with data from the *Régie de l’assurance maladie du Québec* (RAMQ), including fee-for-service physician claims, diagnostic and intervention codes, and patients’ demographic information. We identified children and adolescents aged 0–19 years with at least one diagnostic code for a malignant tumour between January 2015 and December 2023 recorded in MED-ECHO. We categorized cancer diagnoses into 12 diagnostic groups as defined by the International Classification of Childhood Cancer, third edition (ICCC-3).^23^ International Classification of Diseases for Oncology, third edition (ICD-O-3) codes were mapped to ICCC-3 categories using the published conversion algorithm.^23^ We applied a five-year washout period (2010–2014) to exclude prevalent cancer cases and their associated treatments. This secondary data analysis was approved by the McGill Institutional Review Board, in accordance with the Canadian Tri-Council Policy Statement of Ethical Conduct for Research Involving Humans.

### Outcomes

We assessed the monthly age-standardized rates (ASRs) per 1,000,000 population for newly diagnosed cancers, administered chemotherapy, radiotherapy-related services, treatment-related surgeries, positron emission tomography (PET) scans, cancer-related hospital admissions, and all treatments combined. All treatment and healthcare utilization outcomes were assessed among the identified cancer cases only. PET scan utilization was examined as a potential indicator of disruptions to cancer care given its role in disease staging and treatment response assessment.^24,25^ We assigned the date of cancer diagnosis using a previously described hierarchical algorithm that prioritized biopsy as the most definitive diagnostic procedure.^26^ Chemotherapy administration, radiotherapy-related services, treatment-related surgeries, and PET scans were identified using Canadian Classification of Health Interventions (CCI) and physician billing codes. As the setting of treatment delivery could not be reliably determined from physician billing codes, inpatient and outpatient services were not differentiated. Chemotherapy administration was restricted to services provided by specialists in hematology, medical oncology, or pediatric hematology-oncology. Radiotherapy-related services included treatment planning or new radiation services. We compiled billing codes for administered chemotherapy, radiation-related services, and surgery to create a composite measure of all treatments combined, allowing patients to contribute to more than one treatment type within the same month. We assessed all treatments combined overall and stratified by age group for children aged 0-14 years and adolescents aged 15-19 years. Codes used to define each outcome are reported in Supplementary Table 1.

We calculated hospital length of stay as the number of days between admission and discharge. When admission and discharge occurred on the same day, such as in the case of ambulatory surgery, we assigned a length of stay of 0.5 days to reflect partial-day hospital admission.

### Exposures and covariates

We defined the COVID-19 post-pandemic period as March 2020 through end of follow-up to December 2023, beginning with the declaration of the public health emergency in Québec and the implementation of public health restrictions.^27^ January 2015 through February 2020 constituted the pre-pandemic baseline period.

Clinical and demographic characteristics relevant to healthcare access and patterns of care were summarized descriptively, including age at diagnosis (<1, 1–4, 5–9, 10–14, 15–19), cancer type, sex, and quintile of material and social deprivation index, assigned at the census dissemination area level using postal code of residence at diagnosis.^28,29^ We assessed two geography-based variables. Residential rurality was classified based on the patient’s place of residence at diagnosis using Statistics Canada Census Metropolitan Area classification. Health region type, reflecting the geographical location of the facility providing care, was assessed at diagnosis and for each subsequent hospital admission. The health region recorded for each encounter was then categorized into types according to the presence of and proximity to academic and specialized care facilities.^30^

### Statistical analysis

We used an interrupted time series approach to evaluate immediate and sustained changes in childhood and adolescent cancer diagnosis and cancer-related service utilization attributable to the COVID-19 pandemic. We fitted autoregressive integrated moving average (ARIMA) models to monthly ASRs, calculated using direct standardization to the 2021 Canadian standard population aged 0-19 years to account for changes in population size and age structure over time.^31,32^ ARIMA models accounted for autocorrelation, underlying temporal trends, and seasonality in monthly rates. We divided the time series into a pre-pandemic baseline period (January 2015 to February 2020) and a post-pandemic period (March 2020 to December 2023), with March 2020 representing the interruption point. The impact model included a level shift to estimate the immediate change in monthly ASR at the onset of the pandemic, and slope change to estimate any subsequent change in trend relative to the pre-pandemic period.^33^ We assessed variance stability and stationarity using visual inspection of the time series, autocorrelation plots, and the augmented Dickey-Fuller and Kwiatkowski-Phillips-Schmidt-Shin tests.^34,35^ When necessary, we applied nonseasonal or seasonal differencing to achieve stationarity. Candidate models were selected based on autocorrelation and partial autocorrelation function plots and the minimum Akaike information criterion. Residual autocorrelation was assessed using the Ljung-Box test.^32,36,37^ We used each outcome-specific ARIMA model fitted to pre-pandemic data to forecast and plot counterfactual monthly ASRs and corresponding 95% prediction intervals (PI) for the post-pandemic period.

To quantify cumulative differences in the number of cancer diagnoses, chemotherapy administrations, surgeries, radiation services, and all treatments combined among children in Québec, we repeated ARIMA analyses fitted to pre-pandemic monthly event counts rather than ASRs. We generated 10,000 simulated counterfactual trajectories from the fitted models, incorporating both parameter uncertainty through the variance-covariance matrix and forecast uncertainty from future ARIMA innovations. For each simulated trajectory, we summed the observed and expected monthly counts over the early pandemic period (3 months, March 2020–May 2020), mid-pandemic period (12 months, March 2020–February 2021), and total pandemic period (46 months, March 2020–December 2023). We then calculated the cumulative difference by subtracting the expected total from the observed total, and the cumulative ratio by dividing the observed total by the expected total. We used the 2.5^th^ and 97.5^th^ percentiles of the simulated differences and ratios to construct 95% uncertainty intervals (UI). These differences represent the estimated excess or deficit in diagnoses and services relative to counterfactual expectations over specified periods of the pandemic.

We analyzed length of hospital stay at the individual admission level. We restricted to cancer-related hospital admissions occurring after diagnosis among individuals aged 0–19 years identified in the MED-ECHO tumour file between January 2015 and December 2023. We summarized mean length of hospital stay by patient characteristics and stratified by pandemic period. Because length of stay was right-skewed, we fitted univariate and multivariable generalized linear mixed models with a gamma distribution and log link function to estimate associations with patient characteristics.^38^ All models included a random intercept for patient identifier to account for multiple admissions within patients. We adjusted for sex, cancer type, age at hospital admission, pandemic period at admission, material deprivation quintile at admission, health region type of the admitting facility, day of the week, month of admission, and calendar time. For covariates with missing data, we included a missing-indicator category to retain all individuals in the analysis. Model-based marginal mean length of stay was estimated using least-squares means. For selected covariates, we used pairwise least-squares mean contrasts to estimate differences in mean length of stay on the absolute scale (days) and the multiplicative scale (ratios). We calculated confidence intervals for absolute differences using the delta method. To assess whether the association between pandemic period and length of hospital stay differed across patient and clinical characteristics, we fitted separate models that included interaction terms between pandemic period and each of the covariates of interest: sex, cancer type, health region type of the admitting facility, age at admission, and material deprivation quintile. For each multivariable model, we assessed the global interaction using a Wald test. We calculated marginal mean length of stay and estimated differences on the absolute and multiplicative scales for the post-pandemic period compared with the pre-pandemic period within each level of the interacting variable.

## Data availability

This study used data from the © Government of Québec (2022) under licence from the *Institut de la statistique du Québec*. Restrictions apply to access these data, which are regulated by Québec law. Eligible researchers can request access via application through Data Access Services.^39^ Analysis codes are available on the Borealis repository at https://doi.org/10.5683/SP4/VRPWOT.

## RESULTS

We identified 2570 children and adolescents aged 0-19 years diagnosed with cancer in Québec, Canada, between 2015 and 2023, including 1475 cases diagnosed during the pre-pandemic period (January 2015–February 2020) and 1095 during the post-pandemic period (March 2020–December 2023). The demographic, clinical, and geographic characteristics of cancer cases were similar in both periods (Table 1). Overall, leukemias, lymphomas, and CNS tumours predominated, accounting for over half of all diagnoses. Most patients resided in Montréal and were diagnosed in facilities located in university-affiliated health regions. The distribution of dissemination area-level material and social deprivation was similar across time periods, with the largest proportion of pre-pandemic cases occurring in the least socially deprived quintile. Missingness was limited for both material and social deprivation before (6.46%) and during (6.85%) the pandemic.

**Table 1.** Characteristics of children and adolescents diagnosed with cancer in Quebec, Canada, overall and by period.

| Characteristic |  | Overall | Pre-pandemic | Post-pandemic |
|---|---|---|---|---|
|  |  | January 2015 – December 2023 | January 2015 – February 2020 | March 2020 – December 2023 |
|  |  | 2570 (100)<sup>a</sup> | 1475 (57.4) | 1095 (42.6) |
| Sex |  |  |  |  |
|  | Male | 1375 (53.5) | 785 (53.2) | 590 (53.9) |
|  | Female | 1195 (46.5) | 690 (46.8) | 505 (46.1) |
| Age at diagnosis, years |  |  |  |  |
|  | <1 | 180 (7.0) | 100 (6.8) | 75 (6.8) |
|  | 1–4 | 645 (25.1) | 385 (26.2) | 260 (23.7) |
|  | 5–9 | 520 (20.2) | 295 (20.1) | 225 (20.5) |
|  | 10–14 | 550 (21.4) | 300 (20.4) | 245 (22.4) |
|  | 15–19 | 680 (26.5) | 390 (26.5) | 290 (26.5) |
| Cancer type |  |  |  |  |
|  | Leukemia | 655 (25.5) | 355 (24.1) | 305 (27.9) |
|  | Lymphoma | 400 (15.6) | 220 (14.9) | 180 (16.4) |
|  | CNS | 455 (17.7) | 270 (18.3) | 190 (17.4) |
|  | Neuroblastoma | 150 (5.8) | 95 (6.4) | 55 (5.0) |
|  | Retinoblastoma | 45 (1.8) | 25 (1.7) | 20 (1.8) |
|  | Renal tumour | 110 (4.3) | 60 (4.1) | 50 (4.6) |
|  | Hepatic tumour | 40 (1.6) | 20 (1.4) | 20 (1.8) |
|  | Bone tumour | 155 (6.0) | 100 (6.8) | 50 (4.6) |
|  | Soft tissue sarcoma | 150 (5.9) | 95 (6.4) | 55 (5.0) |
|  | Germ cell | 110 (4.3) | 60 (4.1) | 50 (4.6) |
|  | Carcinoma | 285 (11.1) | 170 (11.5)<sup>b</sup> | 125 (11.4)<sup>b</sup> |
|  | Other | 10 (0.4) | – | – |
| Rurality |  |  |  |  |
|  | Montreal (large urban centre, 4M inhabitants) | 1280 (49.9) | 750 (50.6) | 530 (48.6) |
|  | Other large urban centres (>100,000 inhabitants) | 495 (19.3) | 275 (18.6) | 220 (20.2) |
|  | Regional areas (10,000-100,000 inhabitants) | 225 (8.7) | 135 (9.1) | 90 (8.3) |
|  | Small towns/rural (<10,000 inhabitants) | 475 (18.5) | 270 (18.2) | 210 (19.3) |
|  | Missing | 90 (3.5) | 50 (3.4) | 40 (3.7) |
| Health region type at diagnosis |  |  |  |  |
|  | University-affiliated | 2355 (91.6) | 1350 (91.5) | 1005 (91.8) |
|  | Peripheral | 100 (3.9) | 55 (3.7) | 45 (4.1) |
|  | Intermediate, remote, or northern | 115 (4.5) | 70 (4.7) | 45 (4.1) |
| Material Deprivation Index, Quintile |  |  |  |  |
|  | 1 – least | 495 (19.3) | 290 (19.7) | 205 (18.7) |
|  | 2 | 495 (19.3) | 285 (19.4) | 210 (19.2) |
|  | 3 | 485 (18.9) | 270 (18.4) | 215 (19.6) |
|  | 4 | 470 (18.3) | 255 (17.4) | 215 (19.6) |
|  | 5 – most | 455 (17.7) | 275 (18.7) | 175 (15.9) |
|  | Missing | 170 (6.6) | 95 (6.5) | 75 (6.8) |
| Social Deprivation Index, Quintile |  |  |  |  |
|  | 1 – least | 570 (22.2) | 340 (23.1) | 230 (20.9) |
|  | 2 | 495 (19.3) | 270 (18.4) | 225 (20.4) |
|  | 3 | 505 (19.7) | 280 (19.0) | 225 (20.4) |
|  | 4 | 440 (17.1) | 245 (16.7) | 195 (17.7) |
|  | 5 – most | 390 (15.2) | 240 (16.3) | 150 (13.6) |
|  | Missing | 170 (6.6) | 95 (6.5) | 75 (6.8) |
|  |  | Median (IQR) | Median (IQR) | Median (IQR) |
| Length of hospital admission (days) |  | 2.0 (0.5 to 5.0) | 2.0 (0.5 to 5.0) | 2.0 (1.0 to 5.0) |

Abbreviations: CNS, central nervous system tumour; IQR, Interquartile range.
Values may not add up to 100 percent due to random rounding to ensure confidentiality.
<sup>a</sup> Cell counts were rounded to the nearest 5 in accordance with confidentiality requirements. Percentages may not sum to 100% due to rounding.
<sup>b</sup> Categories for Carcinomas and Other cancers were collapsed to prevent disclosure of small cell sizes.

There was no evidence of an immediate level change at the onset of the pandemic or a subsequent change in monthly trend for newly diagnosed cancers, chemotherapy administration, treatment-related surgeries, radiotherapy-related services, all treatments combined, PET scans, or hospital admissions. Estimates were generally imprecise across outcomes, as reflected by wide confidence intervals. For newly diagnosed cancers, the estimated level change in monthly ASR at pandemic onset was 0.80 cases per 1,000,000 population (95% CI: -0.90 to 2.44), and the estimated slope change was -0.05 cases per 1,000,000 population per month (95% CI: -0.10 to 0.01). For all treatments combined, the estimated level change in monthly ASR at pandemic onset was -5.84 treatments per 1,000,000 population (95% CI: -61.6 to 49.9), and the estimated slope change was -1.16 treatments per 1,000,000 per month (95% CI: -3.34 to 1.02). Outcome-specific ARIMA orders and model estimates are reported in Table 2, and observed and counterfactual ASR trends are shown in Figure 1.

**Figure 1.**
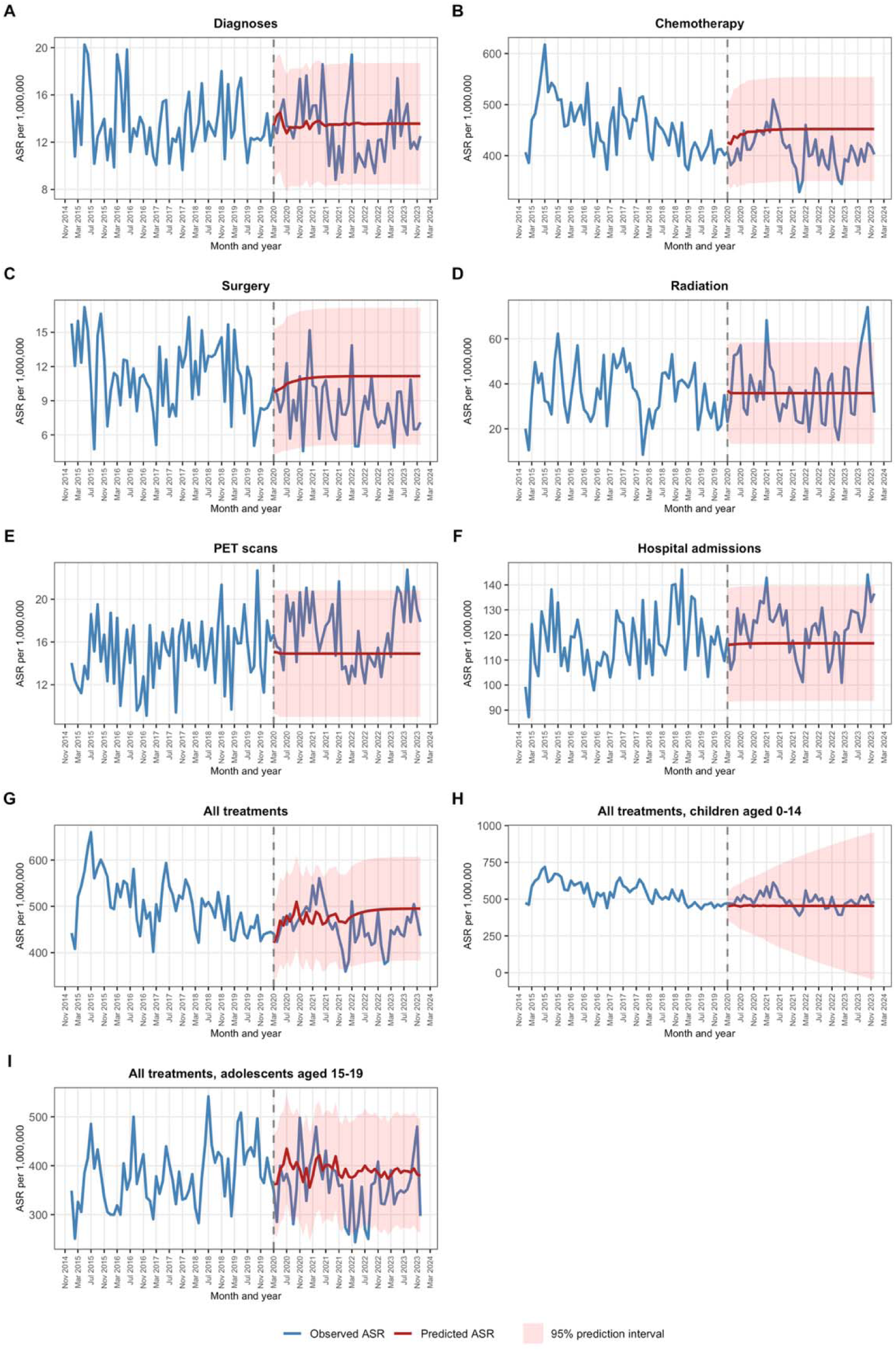
Observed and counterfactual projections of monthly age-standardized rates of cancer diagnosis and treatment-related care: (A) all cancer diagnoses, (B) chemotherapy, (C) cancer-related surgery, (D) radiotherapy-related services, (E) positron emission tomography scans, (F) cancer-related hospital admissions, (G) all treatments combined, (H) all treatments combined among children aged 0–14, and (I) all treatments combined among adolescents aged 15–19.

**Table 2.** ARIMA estimates of pandemic-associated level and slope changes in monthly age-standardized rates of cancer diagnosis and cancer-related service utilization.

| Outcome | ARIMA<sup>a</sup> | Immediate change (Level) | Post-interruption trend change (Slope difference) |
|---|---|---|---|
|  | (p,d,q)(P,D,Q)<sub>12</sub> | ASR difference per 1,000,000 population (95% CI) | Change in monthly ASR per 1,000,000 population (95% CI) |
| Cancer diagnoses |  |  |  |
|  | (0,0,0)(1,0,0) | 0.80 (-0.90 to 2.44) | -0.05 (-0.10 to 0.01) |
| Administered chemotherapies |  |  |  |
|  | (3,0,1)(1,0,0) | -20.5 (-73.5 to 32.6) | -0.82 (-2.80 to 1.15) |
| Surgeries |  |  |  |
|  | (4,0,0)(0,0,0) | -1.20 (-3.52 to 1.20) | -0.06 (-0.14 to 0.02) |
| Radiation-related services |  |  |  |
|  | (0,0,1)(0,0,0) | -0.93 (-11.2 to 9.3) | 0.09 (-0.26 to 0.44) |
| All treatments combined |  |  |  |
| Overall | (1,0,1)(0,0,2) | -5.84 (-61.6 to 49.9) | -1.16 (-3.34 to 1.02) |
| 0–14 years | (0,1,1)(1,0,0) | 10.3 (-71.5 to 92.1) | —<sup>b</sup> |
| 15–19 years | (1,0,0)(2,0,0) | -14.1 (-62.0 to 33.8) | -0.68 (-2.30 to 0.94) |
| PET scans |  |  |  |
|  | (2,0,0)(0,0,0) | 1.24 (-1.21 to 3.70) | 0.03 (-0.06 to 0.11) |
| Hospital admissions |  |  |  |
|  | (1,0,1)(0,0,0) | 2.20 (-8.84 to 13.2) | 0.16 (-0.23 to 0.56) |

Abbreviations: ARIMA, autoregressive integrated moving averages; ASR, age standardized incidence rate; CI, confidence interval; PET, positron emission tomography
<sup>a</sup> ARIMA model orders are presented as ARIMA (p,d,q)(P,D,Q)<sub>12,</sub> where p and P denote nonseasonal and seasonal autoregressive orders; d and D denote nonseasonal and seasonal orders of differencing; and q and Q denote nonseasonal and seasonal moving-average orders. Nonseasonal differencing subtracts the preceding observation from the current observation, whereas seasonal differencing subtracts the observation from the corresponding month in the preceding year to induce stationarity.
<sup>b</sup> Slope change was not estimated for differenced models because differencing analyzes changes between observations rather than the underlying monthly ASR trend.

Table 3 presents the cumulative observed-minus-expected differences and ratios for cancer diagnoses, chemotherapy administration, treatment-related surgeries, radiotherapy-related services, and all treatments combined over the first three months (March–May 2020), 12 months (March 2020–February 2021), and 46 months (March 2020–December 2023) of the pandemic. Across outcomes and time periods, cumulative differences were generally small and compatible with expected variation. For cancer diagnoses, the difference between observed and expected cases was -1 (95% UI: -17 to 15) in the first three months, 24 (95% UI: -9 to 57) during the first 12 months, and 2 (95% UI: -94 to 96) over the full 46-month pandemic period.

**Table 3.** Observed and expected cumulative cancer diagnoses and cancer-related service use across the post-pandemic period.

| Outcome | **March 2020 – May 2020 (3 months)** |  |  |  | **March 2020 – February 2021 (12 months)** |  |  |  | **March 2020 – December 2023 (46 months)** |  |  |  |
|---|---|---|---|---|---|---|---|---|---|---|---|---|
|  | Obs. | Expected<br>(95% CI) | Difference<br>(95% UI) | Ratio<br>(95% UI) | Obs. | Expected<br>(95% CI) | Difference<br>(95% UI) | Ratio<br>(95% UI) | Obs. | Expected<br>(95% CI) | Difference<br>(95% UI) | Ratio<br>(95% UI) |
| **Cancer diagnoses** |  |  |  |  |  |  |  |  |  |  |  |  |
|  | 75 | 75<br>(47 to 101) | -1<br>(-17 to 15) | 0.98<br>(0.81 to 1.26) | 310 | 285<br>(177 to 392) | 24<br>(-9 to 57) | 1.08<br>(0.97 to 1.22) | 1,095 | 1,095<br>(672 to 1,513) | 2<br>(-94 to 96) | 1.00<br>(0.92 to 1.10) |
| **Administered chemotherapies** |  |  |  |  |  |  |  |  |  |  |  |  |
|  | 2,130 | 2,230<br>(1,733 to 2,720) | -103<br>(-499 to 300) | 0.95<br>(0.81 to 1.16) | 9,070 | 8,995<br>(6,822 to 11,173) | 75<br>(-1,408 to 1,636) | 1.01<br>(0.87 to 1.22) | 34,315 | 34,570<br>(25,060 to 44,080) | -254<br>(-7,289 to 7,245) | 0.99<br>(0.83 to 1.27) |
| **Surgeries** |  |  |  |  |  |  |  |  |  |  |  |  |
|  | 50 | 45<br>(17 to 73) | 4<br>(-15 to 23) | 1.09<br>(0.77 to 1.85) | 200 | 215<br>(85 to 342) | -15<br>(-83 to 63) | 0.93<br>(0.71 to 1.47) | 695 | 885<br>(355 to 1,415) | -188<br>(-422 to 98) | 0.79<br>(0.62 to 1.16) |
| **Radiation-related services** |  |  |  |  |  |  |  |  |  |  |  |  |
|  | 195 | 190<br>(74 to 306) | 7<br>(-77 to 92) | 1.04<br>(0.72 to 1.87) | 840 | 750<br>(276 to 1,226) | 91<br>(-106 to 286) | 1.12<br>(0.88 to 1.52) | 3,135 | 2,870<br>(1,039 to 4,700) | 267<br>(-207 to 722) | 1.09<br>(0.94 to 1.30) |
| **All treatments combined** |  |  |  |  |  |  |  |  |  |  |  |  |
|  | 2,375 | 2,470<br>(1,937 to 3,002) | -95<br>(-527 to 353) | 0.96<br>(0.82 to 1.17) | 10,115 | 9,950<br>(7,582 to 12,319) | 162<br>(-1,419 to 1,883) | 1.02<br>(0.88 to 1.23) | 38,150 | 38,225<br>(27,977 to 48,477) | -79<br>(-7,231 to 8,490) | 1.00<br>(0.84 to 1.29) |

Abbreviations: Obs, observed; CI, confidence interval; UI, uncertainty interval
Observed and expected counts were rounded in accordance with confidentiality requirements; consequently, derived differences may not correspond exactly to the displayed values. Expected counts were estimated using pre-pandemic ARIMA models. Differences represent the cumulative observed minus expected counts; ratios represent observed divided by expected counts over each period.

Length of cancer-related hospital stay was right-skewed overall and within each pandemic period, with most admissions ending in same-day discharge (Figure 2). Among the 220 COVID-19-related hospital admissions, 85 listed COVID-19 as the primary reason for admission and 135 as a secondary reason. Mean length-of-stay ratios from univariate and multivariable models are reported in Supplementary Table 2, with associations varying across covariates and their levels. Unadjusted models indicated shorter mean length of stay during the post-pandemic period [ratio, 0.83 (0.79 to 0.88)] compared to the pre-pandemic period, although no difference was observed after adjustment [ratio, 1.01 (0.94 to 1.10)]. Table 4 presents observed mean length of stay before and during the pandemic, together with pandemic-period contrasts from interaction models on the absolute and multiplicative scales. In multivariable models, global Wald tests indicated evidence of interaction between pandemic period and cancer type (p<0.001), health region type (p=0.001), and quintile of material deprivation (p<0.001), but not by sex (p=0.29) or age at admission (p=0.05). Among individuals with retinoblastoma, mean length of stay was shorter in the post-pandemic period, with an adjusted marginal mean difference of -0.43 days (95% CI: -0.70 to -0.16), corresponding to a 35% reduction in mean length of stay [mean length-of-stay ratio 0.65 (95% CI: 0.51 to 0.83)]. Mean length of stay was also shorter among individuals in the fourth quintile of material deprivation [-1.18 days (95% CI: -1.78 to -0.58)], representing a 23% reduction in mean length of stay [mean length-of-stay ratio 0.77 (95% CI: 0.68 to 0.88)]. No differences were observed among individuals in other deprivation quintiles. In contrast, mean length of stay was 42% longer in the post-pandemic period among individuals with bone tumours [+1.62 days (95% CI: 0.77 to 2.46); ratio, 1.42 (95% CI: 1.20 to 1.67)] and 74% longer among admissions to facilities in peripheral healthcare regions [+1.23 days (95% CI: 0.36 to 2.09); ratio, 1.74 (95% CI: 1.23 to 2.46)].

**Figure 2.**
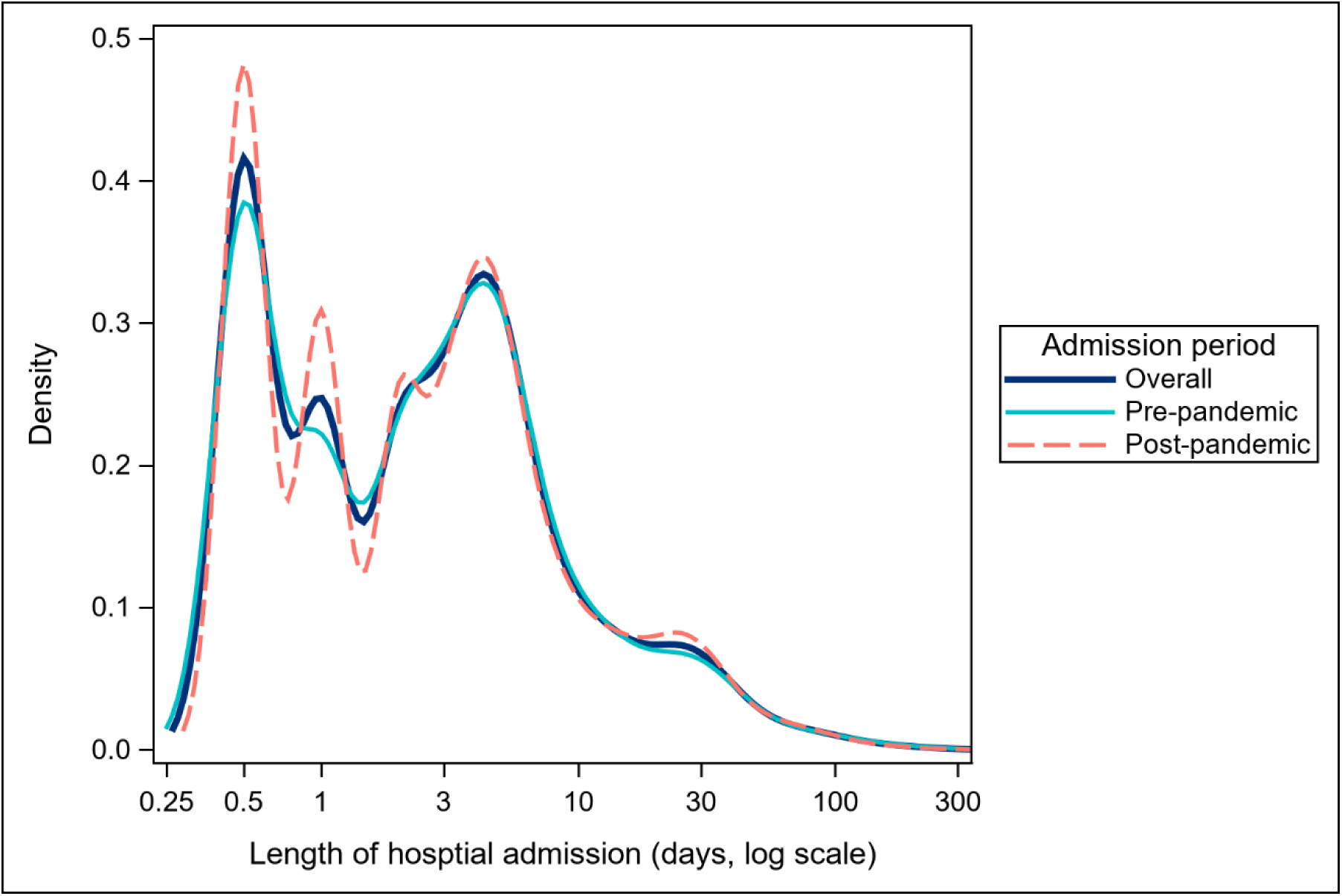
Distribution of length of hospital admission (log scale), overall and by pandemic period. Kernel density plots show the distribution of length of stay (days) on a logarithmic scale for all admissions overall and during the pre- (January 2015–February 2020) and post-pandemic (March 2020–December 2023) periods.

**Table 4.**
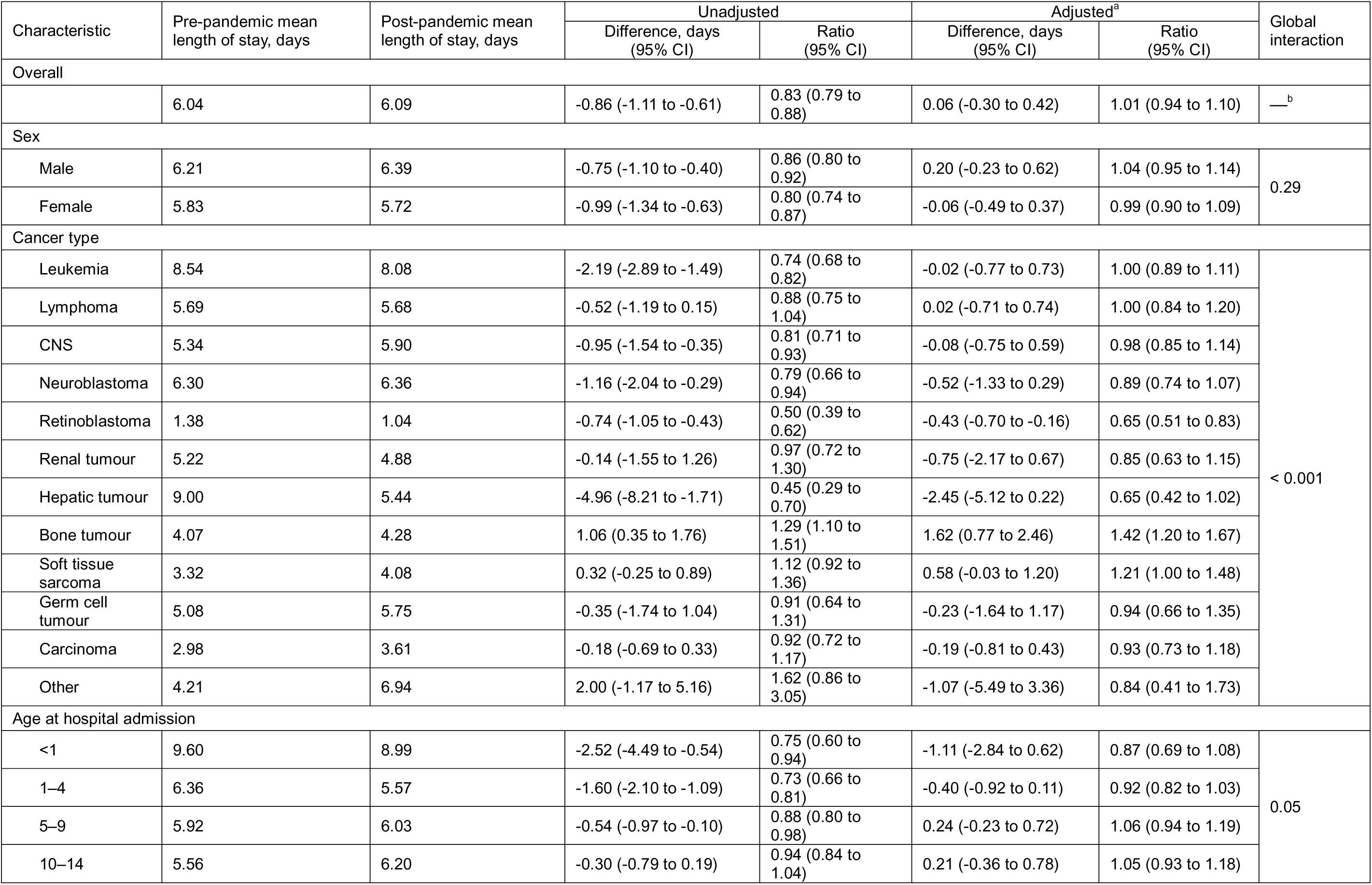
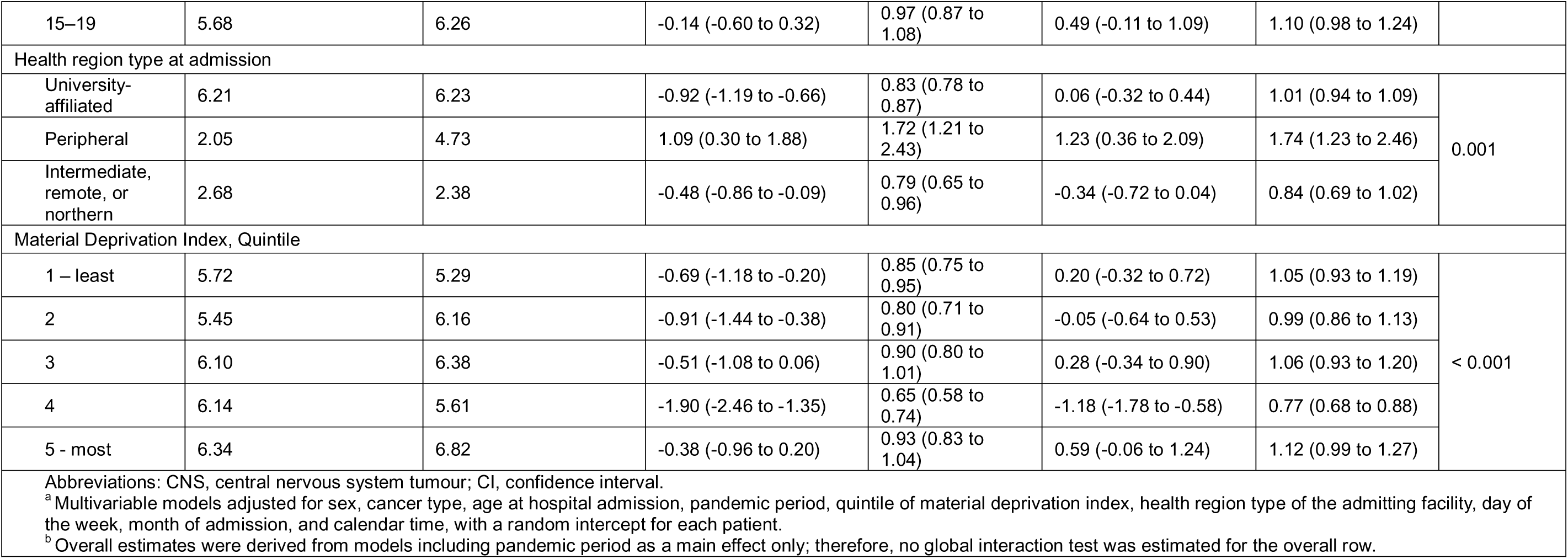
Mean length of hospital stay and pandemic-period contrasts from interaction models, by patient and health system characteristics.

| Characteristic | Pre-pandemic mean length of stay, days | Post-pandemic mean length of stay, days | Unadjusted |  | Adjusted<sup>a</sup> |  | Global interaction |
|---|---|---|---|---|---|---|---|
|  |  |  | Difference, days (95% CI) | Ratio (95% CI) | Difference, days (95% CI) | Ratio (95% CI) |  |
| Overall |  |  |  |  |  |  |  |
|  | 6.04 | 6.09 | -0.86 (-1.11 to -0.61) | 0.83 (0.79 to 0.88) | 0.06 (-0.30 to 0.42) | 1.01 (0.94 to 1.10) | —<sup>b</sup> |
| Sex |  |  |  |  |  |  |  |
| Male | 6.21 | 6.39 | -0.75 (-1.10 to -0.40) | 0.86 (0.80 to 0.92) | 0.20 (-0.23 to 0.62) | 1.04 (0.95 to 1.14) | 0.29 |
| Female | 5.83 | 5.72 | -0.99 (-1.34 to -0.63) | 0.80 (0.74 to 0.87) | -0.06 (-0.49 to 0.37) | 0.99 (0.90 to 1.09) |  |
| Cancer type |  |  |  |  |  |  |  |
| Leukemia | 8.54 | 8.08 | -2.19 (-2.89 to -1.49) | 0.74 (0.68 to 0.82) | -0.02 (-0.77 to 0.73) | 1.00 (0.89 to 1.11) | < 0.001 |
| Lymphoma | 5.69 | 5.68 | -0.52 (-1.19 to 0.15) | 0.88 (0.75 to 1.04) | 0.02 (-0.71 to 0.74) | 1.00 (0.84 to 1.20) |  |
| CNS | 5.34 | 5.90 | -0.95 (-1.54 to -0.35) | 0.81 (0.71 to 0.93) | -0.08 (-0.75 to 0.59) | 0.98 (0.85 to 1.14) |  |
| Neuroblastoma | 6.30 | 6.36 | -1.16 (-2.04 to -0.29) | 0.79 (0.66 to 0.94) | -0.52 (-1.33 to 0.29) | 0.89 (0.74 to 1.07) |  |
| Retinoblastoma | 1.38 | 1.04 | -0.74 (-1.05 to -0.43) | 0.50 (0.39 to 0.62) | -0.43 (-0.70 to -0.16) | 0.65 (0.51 to 0.83) |  |
| Renal tumour | 5.22 | 4.88 | -0.14 (-1.55 to 1.26) | 0.97 (0.72 to 1.30) | -0.75 (-2.17 to 0.67) | 0.85 (0.63 to 1.15) |  |
| Hepatic tumour | 9.00 | 5.44 | -4.96 (-8.21 to -1.71) | 0.45 (0.29 to 0.70) | -2.45 (-5.12 to 0.22) | 0.65 (0.42 to 1.02) |  |
| Bone tumour | 4.07 | 4.28 | 1.06 (0.35 to 1.76) | 1.29 (1.10 to 1.51) | 1.62 (0.77 to 2.46) | 1.42 (1.20 to 1.67) |  |
| Soft tissue sarcoma | 3.32 | 4.08 | 0.32 (-0.25 to 0.89) | 1.12 (0.92 to 1.36) | 0.58 (-0.03 to 1.20) | 1.21 (1.00 to 1.48) |  |
| Germ cell tumour | 5.08 | 5.75 | -0.35 (-1.74 to 1.04) | 0.91 (0.64 to 1.31) | -0.23 (-1.64 to 1.17) | 0.94 (0.66 to 1.35) |  |
| Carcinoma | 2.98 | 3.61 | -0.18 (-0.69 to 0.33) | 0.92 (0.72 to 1.17) | -0.19 (-0.81 to 0.43) | 0.93 (0.73 to 1.18) |  |
| Other | 4.21 | 6.94 | 2.00 (-1.17 to 5.16) | 1.62 (0.86 to 3.05) | -1.07 (-5.49 to 3.36) | 0.84 (0.41 to 1.73) |  |
| Age at hospital admission |  |  |  |  |  |  |  |
| <1 | 9.60 | 8.99 | -2.52 (-4.49 to -0.54) | 0.75 (0.60 to 0.94) | -1.11 (-2.84 to 0.62) | 0.87 (0.69 to 1.08) | 0.05 |
| 1–4 | 6.36 | 5.57 | -1.60 (-2.10 to -1.09) | 0.73 (0.66 to 0.81) | -0.40 (-0.92 to 0.11) | 0.92 (0.82 to 1.03) |  |
| 5–9 | 5.92 | 6.03 | -0.54 (-0.97 to -0.10) | 0.88 (0.80 to 0.98) | 0.24 (-0.23 to 0.72) | 1.06 (0.94 to 1.19) |  |
| 10–14 | 5.56 | 6.20 | -0.30 (-0.79 to 0.19) | 0.94 (0.84 to 1.04) | 0.21 (-0.36 to 0.78) | 1.05 (0.93 to 1.18) |  |
| 15–19 | 5.68 | 6.26 | -0.14 (-0.60 to 0.32) | 0.97 (0.87 to 1.08) | 0.49 (-0.11 to 1.09) | 1.10 (0.98 to 1.24) |  |
| Health region type at admission |  |  |  |  |  |  |  |
| University-affiliated | 6.21 | 6.23 | -0.92 (-1.19 to -0.66) | 0.83 (0.78 to 0.87) | 0.06 (-0.32 to 0.44) | 1.01 (0.94 to 1.09) | 0.001 |
| Peripheral | 2.05 | 4.73 | 1.09 (0.30 to 1.88) | 1.72 (1.21 to 2.43) | 1.23 (0.36 to 2.09) | 1.74 (1.23 to 2.46) |  |
| Intermediate, remote, or northern | 2.68 | 2.38 | -0.48 (-0.86 to -0.09) | 0.79 (0.65 to 0.96) | -0.34 (-0.72 to 0.04) | 0.84 (0.69 to 1.02) |  |
| Material Deprivation Index, Quintile |  |  |  |  |  |  |  |
| 1 – least | 5.72 | 5.29 | -0.69 (-1.18 to -0.20) | 0.85 (0.75 to 0.95) | 0.20 (-0.32 to 0.72) | 1.05 (0.93 to 1.19) | < 0.001 |
| 2 | 5.45 | 6.16 | -0.91 (-1.44 to -0.38) | 0.80 (0.71 to 0.91) | -0.05 (-0.64 to 0.53) | 0.99 (0.86 to 1.13) |  |
| 3 | 6.10 | 6.38 | -0.51 (-1.08 to 0.06) | 0.90 (0.80 to 1.01) | 0.28 (-0.34 to 0.90) | 1.06 (0.93 to 1.20) |  |
| 4 | 6.14 | 5.61 | -1.90 (-2.46 to -1.35) | 0.65 (0.58 to 0.74) | -1.18 (-1.78 to -0.58) | 0.77 (0.68 to 0.88) |  |
| 5 - most | 6.34 | 6.82 | -0.38 (-0.96 to 0.20) | 0.93 (0.83 to 1.04) | 0.59 (-0.06 to 1.24) | 1.12 (0.99 to 1.27) |  |

Abbreviations: CNS, central nervous system tumour; CI, confidence interval.
<sup>a</sup> Multivariable models adjusted for sex, cancer type, age at hospital admission, pandemic period, quintile of material deprivation index, health region type of the admitting facility, day of the week, month of admission, and calendar time, with a random intercept for each patient.
<sup>b</sup> Overall estimates were derived from models including pandemic period as a main effect only; therefore, no global interaction test was estimated for the overall row.

## DISCUSSION

In this population-based study of children and adolescents with cancer in Québec, Canada, we found no evidence of immediate or sustained changes in cancer diagnosis or cancer-related service utilization following the onset of the COVID-19 pandemic. Monthly variations in diagnoses and treatments were largely within expected ranges given pre-pandemic trends. We also found no evidence of cumulative differences between observed and expected numbers of diagnoses or treatments over the first 3, 12, or full 46 months following pandemic onset. However, we did observe changes in length of hospital stay from March 2020 onward for select cancer types, health region types, and quintiles of material deprivation.

Our findings are consistent with earlier pan-Canadian evidence suggesting no disruption in the diagnosis of childhood cancer during the early pandemic period.^16^ Our cumulative analyses further showed no long-term evidence of excess or fewer diagnoses over the full pandemic period, despite reports of differences during later pandemic years.^7^ However, the extent to which these later-period differences were attributable to the pandemic remains unclear.^17,32,33^ The absence of detectable change in childhood cancer diagnoses contrasts with the 15% reduction in reporting of new adult cancers in Québec during the first year of the pandemic,^8^ and may reflect differences in how cancers come to clinical attention in children. Although encounters with family physicians and emergency departments declined among children across Canada,^14,40,41^ the pediatric emergency department visits that did occur tended to involve higher-acuity presentations.^40^ Because childhood cancer diagnosis is largely symptom-driven rather than reliant on asymptomatic screening, families may have continued to seek care for acute, persistent, or progressively concerning presentations, preserving opportunities for cancer recognition despite lower overall healthcare utilization.

Evidence of pandemic-related disruption was also limited for treatment-related activity, PET scans, and hospital admissions, despite reports of disruptions to cancer services in other settings. In Ontario, early reductions in cancer-related surgery and radiotherapy services occurred alongside increases in systemic treatment and PET scans,^19^ suggesting that pandemic effects on cancer services varied across service types. In contrast, we observed no evidence of immediate or sustained changes in monthly rates of treatment-related activity. While treatment backlogs have been reported, particularly for cancer-related surgeries,^4,19,42^ cumulative treatment activity in our study remained within expected levels over the entire 46-month pandemic period, suggesting that any short-term disruptions did not develop into sustained deficits at the population level.

The absence of observed changes in core pediatric oncology services in Québec may partially reflect provincial guidance and institutional measures intended to prioritize care for children and adolescents with cancer.^43,44^ This prioritization was also reflected in treatment-specific guidance. Although reductions in radiotherapy activity were recommended for the broader patient population, treatment for pediatric patients was to continue according to clinical need despite the risk of COVID-19 infection.^20,21^ Guidance similarly prioritized clinical trial enrolment and participation, reinforcing efforts to minimize pandemic-related departures from planned treatment.^21^ Importantly, provincial guidance was not universally restrictive, with children receiving or recently completing treatment encouraged to return to school in September 2020 when clinically appropriate, and siblings encouraged to attend even when the child with cancer was advised to remain home.^45^ While the relative contribution of individual protective measures was not investigated, the absence of detectable changes in diagnosis, treatment, and hospital admission rates is compatible with a role for provincial guidance in maintaining pediatric oncology services.

While length of hospital stay has important implications for patient morbidity, family well-being, and healthcare utilization, its clinical relevance and interpretation are context dependent.^46^ Interactions between pandemic period and cancer type, health region type, and material deprivation indicated that changes in length of stay were non-uniform in both direction and magnitude. Indeed, pandemic-related changes in care could plausibly have contributed to longer or shorter stays: COVID-19 testing requirements, preoperative admissions, and isolation protocols may have prolonged some hospitalizations,^4,47–50^ while efforts to reduce in-hospital transmission may have favoured earlier discharge when clinically feasible.^18,44^ For patients with bone tumours, whose care frequently involves complex surgical intervention, observed increases in length of stay may have been influenced by pandemic-related changes in perioperative care,^44,49^ including prolonged or earlier hospitalization to facilitate access to constrained operating room capacity.^51^ Conversely, shorter stays among patients with retinoblastoma may reflect earlier discharge following enucleation, a procedure with predictable postoperative recovery.^52,53^ Variation in length of stay by health region type of the admitting facility may also reflect differences in admission practices.^40,41^ In peripheral health regions, characterized by greater geographic separation from major health centres,^30^ brief or precautionary admissions may have been reduced with increased use of virtual care.^54^ In contrast, admissions in university-affiliated health regions, where protocol-driven cancer treatment and complex care are concentrated,^55^ may have been less susceptible to such changes given the prioritization of essential pediatric oncology services. While greater material deprivation has been previously associated with longer hospital admission among children with medical complexity,^56^ we observed no consistent gradient. The isolated reduction in length of stay among those in the fourth deprivation quintile should therefore be interpreted cautiously and may reflect chance variation. More broadly, changes in admission practices and the use of alternative care pathways may have altered the case mix of hospitalized patients,^20,21^ such that observed differences in length of stay partially reflect who was admitted rather than changes in inpatient care itself. Accordingly, the clinical significance of changes in length of stay during the pandemic remains unclear.

### Strengths and limitations

We used administrative data to capture cancer-related healthcare encounters among children and adolescents in Québec, Canada, from 2015 to 2023. These data provide near-complete population coverage and include care delivered at both pediatric and adult facilities, reducing the potential for missed encounters across care settings. However, the available data only permitted identification of malignant diagnoses. Because ICCC-3 includes benign and borderline tumours within the CNS and germ cell tumour diagnostic groups,^23^ some of these tumours were not captured. This is particularly relevant for CNS tumours, for which non-malignant neoplasms comprise approximately 13–18% of all diagnoses in children.^57^ Further, bundled billing codes prevented identification of the specific care delivered during some encounters, while billing frequency restrictions and initiation-only billing codes limited the capture of ongoing treatment encounters, potentially underestimating monthly rates.^58^ Information on treatment route and scheduling was also insufficient to characterize treatment phase, delivery, and intensity, including potential adaptations to treatment delivery during the pandemic; we were unable to reliably differentiate treatments delivered in inpatient versus outpatient settings. The rarity of childhood cancer precluded stratified interrupted time series analyses, potentially obscuring heterogeneity across diagnostic groups. Small monthly counts and variability in rates also limited the precision of our estimates, warranting cautious interpretation of the findings. Monthly aggregation may also have obscured brief delays in diagnosis or treatment occurring within the same calendar month. While length-of-stay analyses accounted for repeated admissions and estimated subgroup differences on both absolute and relative scales, we could not distinguish planned from unplanned admissions, complicating attribution of observed differences to care practices versus clinical circumstances.

## CONCLUSION

Our findings suggest that cancer diagnosis and treatment services for children in Québec, Canada, remained largely stable during the pandemic despite widespread healthcare disruption. While we observed changes in length of hospital stay across select subgroups, the observed pattern of care is consistent with the objectives of governmental and institutional measures to preserve pediatric oncology services during the pandemic.

## Data Integrity

Callum Mullen, Talía Malagón, and Mariam El-Zein had full access to all data and take responsibility for the integrity of the data, the accuracy of the analysis, and reporting.

## Acknowledgements

This work was supported by the Canadian Institutes of Health Research (operating grant VR5-172666 and foundation grant 143347 to Eduardo L. Franco). Callum Mullen is supported by a Doctoral Training Award from the Fonds de recherche du Québec – Santé and a Kuok Fellowship from the Rossy Cancer Network. Talía Malagón is supported by a Junior 1 Salary award from the Fonds de recherche du Québec – Santé. We gratefully acknowledge Sarah Botting-Provost and Rodrigo Noorani from the Division of Cancer Epidemiology at McGill University for their contributions to the development of the codebooks (translation from French to English) and the maintenance of documentation for data used in this study.

## References

1. Siegel RL, Kratzer TB, Wagle NS, Sung H, Jemal A. Cancer statistics, 2026. Ca. 2026;76:e70043.

2. Force LM, Kocarnik JM, May ML, et al. Global burden of cancer in children and adolescents aged 0–19 years, 1990–2023: a systematic analysis for the Global Burden of Disease Study 2023. The Lancet. 2026;407:1360–73.

3. Lam CG, Howard SC, Bouffet E, Pritchard-Jones K. Science and health for all children with cancer. Science. 2019;363:1182–6.

4. Graetz D, Agulnik A, Ranadive R, et al. Global effect of the COVID-19 pandemic on paediatric cancer care: a cross-sectional study. The Lancet Child & Adolescent Health. 2021;5:332–40.

5. Heer E, Ruan Y, Boyne DJ, et al. Impact of the COVID-19 pandemic on cancer diagnoses, stage and survival in Alberta. CMAJ. 2023;195:E804–E12.

6. Decker KM, Feely A, Bucher O, et al. New Cancer Diagnoses Before and During the COVID-19 Pandemic. JAMA Network Open. 2023;6:e2332363-e.

7. Burus T, Lei F, Huang B, et al. Undiagnosed cancer cases in the US during the first 10 months of the COVID-19 pandemic. JAMA oncology. 2024;10:500–7.

8. Ramanakumar AV, Annie B, Frederic L, Christine B, Cathy R, Jean L. Evaluating the impact of COVID-19 on cancer declarations in Quebec, Canada. Cancer Medicine. 2023;12:6260–9.

9. Elek P, Csanádi M, Fadgyas-Freyler P, et al. Heterogeneous impact of the COVID-19 pandemic on lung, colorectal and breast cancer incidence in Hungary: results from time series and panel data models. BMJ open. 2022;12:e061941.

10. Coma E, Guiriguet C, Mora N, et al. Impact of the COVID-19 pandemic and related control measures on cancer diagnosis in Catalonia: a time-series analysis of primary care electronic health records covering about five million people. BMJ open. 2021;11:e047567.

11. Barclay NL, Pineda Moncusí M, Jödicke AM, et al. The impact of the UK COVID-19 lockdown on the screening, diagnostics and incidence of breast, colorectal, lung and prostate cancer in the UK: a population-based cohort study. Frontiers in oncology. 2024;14:1370862.

12. Force LM, Abdollahpour I, Advani SM, et al. The global burden of childhood and adolescent cancer in 2017: an analysis of the Global Burden of Disease Study 2017. The Lancet Oncology. 2019;20:1211–25.

13. Mullen CJR, Barr RD, Franco EL. Timeliness of diagnosis and treatment: the challenge of childhood cancers. Br J Cancer. 2021;125:1612–20.

14. Canadian Institute for Health Information. COVID-19’s impact on physician services. 2021; https://www.cihi.ca/en/covid-19-resources/impact-of-covid-19-on-canadas-health-care-systems/physician-services. Accessed June 30, 2026.

15. Dvori M, Elitzur S, Barg A, et al. Delayed diagnosis and treatment of children with cancer during the COVID-19 pandemic. International journal of clinical oncology. 2021;26:1569–74.

16. Pelland-Marcotte M-C, Xie L, Barber R, et al. Incidence of childhood cancer in Canada during the COVID-19 pandemic. Cmaj. 2021;193:E1798–E806.

17. Xie L, Gibson P, Barber R, et al. Changes in childhood cancer incidence rates and trends in Canada (2001–2023): Results from the Cancer in Young People in Canada surveillance program. Cancer Epidemiology. 2026;103:103106.

18. Tran A, Hou SH, Forbes C, et al. The impact of the early COVID-19 global pandemic on children undergoing active cancer treatment and their parents. Current Oncology. 2023;30:2441–56.

19. Walker MJ, Wang J, Mazuryk J, et al. Delivery of cancer care in Ontario, Canada, during the first year of the COVID-19 pandemic. JAMA Network Open. 2022;5:e228855.

20. Ministère de la Santé et des Services sociaux. Recommandations générales en cancérologie en contexte de la COVID-19. Québec (Québec) Octobre 2020 2020. DGAUMIP-005.

21. Ministère de la Santé et des Services sociaux. Recommandations concernant la prise en charge de la clientèle en oncologie pédiatrique dans le contexte de la COVID. Québec (Québec) 14 avril 2020 2020.

22. Régie de l’assurance maladie. La RAMQ en quelques chiffres. 2024; https://www.ramq.gouv.qc.ca/en/node/89526. Accessed November 24, 2025.

23. Steliarova-Foucher E, Stiller C, Lacour B, Kaatsch P. International Classification of Childhood Cancer, third edition. Cancer. 2005;103:1457–67.

24. McCarville MB. PET-CT imaging in pediatric oncology. Cancer Imaging. 2009;9:35.

25. Bradley NM, Robinson PD, Greenberg ML, et al. Measuring the quality of a childhood cancer care delivery system: quality indicator development. Value in Health. 2013;16:647–54.

26. Mullen C, Barr RD, Strumpf E, El-Zein M, Franco EL, Malagón T. Time to Diagnosis Among Children and Adolescents With Cancer in Québec, Canada: A Population-Based Study. Pediatric Blood & Cancer. 2026:e70610.

27. Gouvernement du Québec. Décret 177-2020 du 13 mars 2020 concernant une déclaration d’urgence sanitaire conformément à l’article 118 de la Loi sur la santé publique. *Gazette officielle du Québec*, Lois et règlements. 2020;152:1101A–2A.

28. Pampalon R, Hamel D, Gamache P, Philibert MD, Raymond G, Simpson A. An Area-based Material and Social Deprivation Index for Public Health in Québec and Canada. Canadian Journal of Public Health / Revue Canadienne de Sante’e Publique. 2012;103:S17–S22.

29. Azevedo da Silva M, Gravel N, Sylvain-Morneau J, Blaser C, Gamache P, Hamel D. Material and Social Deprivation Index 2021: User Manual. Québec, Canada: Institut national de santé publique du Québec; March 2024.

30. Gauthier J, Haggerty J, Lamarche P, et al. Entre adaptabilité et fragilité: les conditions d’accès aux services de santé des communautés rurales et éloignées: Institut national de santé publique du Québec Québec; 2009.

31. Statistics Canada. Canadian Cancer Registry - Standard population used for Age-standardization. 2025; https://www.statcan.gc.ca/en/statistical-programs/document/3207_D12_V5. Accessed June 10, 2026.

32. Schaffer AL, Dobbins TA, Pearson S-A. Interrupted time series analysis using autoregressive integrated moving average (ARIMA) models: a guide for evaluating large-scale health interventions. BMC medical research methodology. 2021;21:58.

33. Bernal JL, Cummins S, Gasparrini A. Interrupted time series regression for the evaluation of public health interventions: a tutorial. International journal of epidemiology. 2017;46:348–55.

34. Dickey DA, Fuller WA. Likelihood ratio statistics for autoregressive time series with a unit root. Econometrica: journal of the Econometric Society. 1981:1057–72.

35. Kwiatkowski D, Phillips PC, Schmidt P, Shin Y. Testing the null hypothesis of stationarity against the alternative of a unit root: How sure are we that economic time series have a unit root? Journal of econometrics. 1992;54:159–78.

36. Box GE, Jenkins GM, Reinsel GC, Ljung GM. Time series analysis: forecasting and control: John Wiley & Sons; 2015.

37. Box GE, Tiao GC. Intervention analysis with applications to economic and environmental problems. Journal of the American Statistical association. 1975;70:70–9.

38. Williford E, Haley V, McNutt L-A, Lazariu V. Dealing with highly skewed hospital length of stay distributions: the use of Gamma mixture models to study delivery hospitalizations. PLoS One. 2020;15:e0231825.

39. Institut de la statistique du Québec. Submitting a request. https://statistique.quebec.ca/en/services-recherche/submitting-a-request. Accessed August 4, 2026.

40. Finkelstein Y, Maguire B, Zemek R, et al. Effect of the COVID-19 pandemic on patient volumes, acuity, and outcomes in pediatric emergency departments: a nationwide study. Pediatric emergency care. 2021;37:427–34.

41. Iskander C, Stukel TA, Diong C, et al. Acute health care use among children during the first 2.5 years of the COVID-19 pandemic in Ontario, Canada: a population-based repeated cross-sectional study. CMAJ. 2024;196:E1–E13.

42. Eskander A, Li Q, Hallet J, et al. Access to cancer surgery in a universal health care system during the COVID-19 pandemic. JAMA network open. 2021;4:e211104.

43. Ministère de la Santé et des Services Sociaux. Directives COVID-19 du Ministère de la Santé et des Services Sociaux. 2020; https://publications.msss.gouv.qc.ca/msss/document-002698/. Accessed November, 2025.

44. Sullivan M, Bouffet E, Rodriguez-Galindo C, et al. The COVID-19 pandemic: a rapid global response for children with cancer from SIOP, COG, SIOP-E, SIOP-PODC, IPSO, PROS, CCI, and St Jude Global. Pediatric blood & cancer. 2020;67:e28409.

45. Ministère de la Santé et des Services sociaux. Recommandations du Comité national de cancérologie pédiatrique en contexte de la pandémie de la COVID-19 – Retour à l’école et à la garderie en septembre 2020. Québec (Québec) 27 August 2020 2020.

46. Cram P. Hospital length of stay….. a measure of what, exactly? Medical care. 2019;57:751.

47. Ban Y, Hoshi M, Oebisu N, et al. Impact of the COVID-19 pandemic on bone and soft tissue tumor treatment: A single-institution study. PLoS One. 2023;18:e0283835.

48. Brunner L, Nicolet A, Peytremann-Bridevaux I, et al. Comparative inpatient care of cancer vs. non-cancer patients in Switzerland during the national COVID-19 lockdown: a nationwide interrupted time series analysis. BMC cancer. 2025;25:477.

49. IWK Health Centre. COVID-19 Pandemic Plan for Pediatric Hematology and Oncology Patients: Maritime Provinces. Halifax, Nova Scotia: IWK Health Centre Pediatric Hematology Oncology Program; May 2021.

50. Sniderman ER, Graetz DE, Agulnik A, et al. Impact of the COVID-19 pandemic on pediatric oncology providers globally: a mixed-methods study. Cancer. 2022;128:1493–502.

51. Bouthillier M-E, Lorange M, Legault S, et al. Prioritizing surgery during the COVID-19 pandemic: the Quebec guidelines. Canadian Journal of Surgery. 2021;64:E103.

52. Skalet AH, Allen RC, Shields CL, Wilson MW, Mruthyunjaya P, Gombos DS. Considerations for the management and triage of ocular oncology cases during the COVID-19 pandemic. Ocular Oncology and Pathology. 2020;6:219–22.

53. Canadian Cancer Society. Surgery for retinoblastoma. 2025; https://cancer.ca/en/cancer-information/cancer-types/retinoblastoma/treatment/surgery. Accessed July 3, 2026.

54. Mitra A, Veerakone R, Li K, Nix T, Hashikawa A, Mahajan P. Telemedicine in paediatric emergency care: a systematic review. Journal of Telemedicine and Telecare. 2023;29:579–90.

55. Mitra D, Hutchings K, Shaw A, et al. The Cancer in Young People in Canada surveillance system. Health Promotion and Chronic Disease Prevention in Canada: Research, Policy and Practice. 2015;35:73.

56. Warniment A, Zhang Y, Huang B, Thomson J, Auger KA. Neighborhood socioeconomic deprivation and length of stay in children with medical complexity. Hospital pediatrics. 2025;15:474–82.

57. Stiller C. Pediatric cancers. International Encyclopedia of Public Health (Second Edition): Elsevier Inc.; 2017:426–37.

58. Konrad R, Zhang W, Bjarndóttir M, Proaño R. Key considerations when using health insurance claims data in advanced data analyses: an experience report. Health Systems. 2020;9:317–25.

