## Supplementary Material for "Childhood and adolescent cancer diagnosis, treatment, and hospitalizations during the COVID-19 pandemic in Québec, Canada, a population-based study"

**Table of Contents**

Supplementary Table S1. Billing codes used to define ARIMA outcomes ..……..……….…. 2–5

Supplementary Table S2. Estimated mean length of stay ratios ……….….……………..……. 6–7

| **Supplementary Table S1.** Billing codes used to define cancer-related outcomes evaluated in ARIMA models | | |
| --- | --- | --- |
|  | ACTE  physician billing codes | Canadian Classification of Health Interventions codes |
| **Chemotherapy** | | |
|  | 00583, 00734, 00603, 00595, 00596 | 1ZZ35HA, 1ZZ35CA, 1ZZ35YA, 1AX35HA |
| **Surgery** | | |
| Lymphoma | 04241, 04159, 04161, 04199, 04234, 04235, 04240, 04242, 04243, 04244, 04245, 04233, 18171, 18172, 18173 |  |
| Central nervous system tumours | 07650, 07000, 07520, 07521, 07522, 07523, 07524, 07525, 07526, 07527, 07528, 07529, 07530, 07532, 07533, 07534, 07535, 07536, 07537, 07538, 07539, 07540, 07541, 07542, 07543, 07545, 07546, 07547, 07548, 07549, 07550, 07551, 07552, 07553, 07554, 07555, 07556, 07652, 07653, 07684, 07685, 07686, 07690, 07691, 07692, 07790, 18171, 18172, 18173 | 1AX87DA, 1AX87LA, 1AA87SZ, 1AB87SE, 1AB87SZ, 1AC87DA, 1AC87SZ, 1AJ87DA, 1AJ87PN, 1AJ87SZ, 1AK87PN, 1AK87PQ, 1AK87QQ, 1AK87SZ, 1AN87PN, 1AN87SE, 1AN87SZ, 1AP87LP, 1AP87PN, 1AP87PP, 1AP87SZ, 1AW87DA, 1AW87LA, 1AX87DA, 1AX87GB, 1AX87LA, 1AX87WK, 1CJ87LA, 1AE87SZ, 1AF87DA, 1AF87LU, 1AF87PR, 1AG87SE, 1AG87SZ, 1AJ87DA, 1AJ87PN, 1AJ87SZ, 1AK87PN, 1AK87PQ, 1AK87QQ, 1AK87SZ, 1AN87PN, 1AN87SE, 1AN87SZ, 1AP87LP, 1AP87PN, 1AW87DA, 1AW87LA, 1AX87DA, 1AX87GB, 1AX87LA, 1AX87WK |
| Neuroblastoma | 05480, 06084, 06161, 06162, 06217, 03196, 05193, 05480, 06161, 06179, 06200, 07172, 07189, 06101, 06182, 18171, 18172, 18173 | 1OT87DA, 1OT87LA, 1OT91LA, 1BA87UH, 1BA87WF, 1BB87LA, 1BB87UH, 1BB87WF, 1BB87WG, 1BD87QQ, 1BD87XE, 1BG87LA, 1BG87UH, 1BG87WF, 1BG87WG,1BM87LA, 1BM87UH, 1BN87LA, 1BN87UH, 1BP87LA, 1BP87UH, 1BQ87LA, 1BS87LA, 1BT87LA, 1BX87LA, 1PB87DA, 1PB87LB, 1PB87PF, 1PB89DA, 1PB89LB, 1PB89PF, 1PB89QF |
| Retinoblastoma | 07310, 07133, 07134, 07311, 18171, 18172, 18173 | 1CP87LA, 1CP87SZ, 1CP87RZ, 1CP91LA, 1CP89LA |
| Renal tumours | 06084, 06111, 06161, 06162, 06217, 03196, 05193, 05480, 06161, 06179, 06200, 06217, 06101, 06182, 18171, 18172, 18173 | 1PC87LA, 1PC91LB, 1PC87DA, 1PC89PF, 1PC89LB |
| Hepatic tumours | 05486, 05480, 05145, 05146, 05147, 05485, 18171, 18172, 18173 | 1OT87DA, 1OT91LA, 1OT87LA, 1OA87DA, 1OA87LA |
| Bone tumours | 02130, 02131, 02132, 02137, 02165, 02167, 02168, 02178, 02179, 02181, 02722, 02723, 02780, 02781, 02787, 02788, 02789, 02790, 02791, 02799, 02808, 02811, 02812, 02813, 02870, 02871, 02995, 02996, 02997, 09509, 09510, 09511, 02215, 09560, 02271, 02273, 02274, 02275, 02278, 02279, 02280, 02281, 02282, 02285, 02287, 02292, 02294, 02295, 02296, 18171, 18172, 18173 | 1SN93LA, 1SQ93LA, 1TV93LA, 1VC93LA, 1VG93LA, 1VQ93LA, 1WA93LA, 1SK87LA, 1SL87LL, 1SL87PF, 1SN87LA, 1SQ87LA, 1TA87DA, 1TA87LA, 1TK87LA, 1TV87LA, 1VA87DA, 1VA87LA, 1VC87LA, 1SL91PF, 1SN91LA, 1SQ91LA, 1TK91LA, 1TV91LA, 1VC91LA, 1VQ91LA, 1SL89LL, 1TX87LA, 1SK87LA, 1SL87LL, 1SL87PF, 1SM87LA, 1SN87LA, 1SQ87LA, 1SY87LA, 1SZ87DA, 1SZ87LA, 1TV87LA, 1TX87LA, 1VC87LA, 1EP87LA, 1EQ87LA, 1EC87LA, 2DL71LA, 1EE87LA, 1EB87LA, 1EA87BA |
| Soft tissue sarcomas | 05480, 02130, 02152, 02153, 02154, 09509, 09510, 09511, 02007, 02083, 02127, 02023, 02112, 02208, 02209, 02082, 18171, 18172, 18173, | 1CP87SZ, 1FA87CA, 1OT87DA, 1OT87LA, 1RS87LA, 1SG87LA, 1TF87LA, 1TQ87LA, 1VR87LA, 1EP87LA, 1EQ87LA, 1FA87BA, 1FA87CA, 1FA91LA, 1SG87LA, 1SH87LA, 1SY87LA, 1SZ87DA, 1SZ87LA, 1TX87LA, 1UY87LA, 1VD87LA, 1VX87LA, 1FB87LA, 1FB87LA, 1FC87LA, 1HB87DA, 1HB87LA, 1VX87LA, 1UY87LA, 1FM89LA, 1FM89VW, 1CP87LA, 1WV87LA, 1EY87LA, 1ES87LA, 2NK71BA, 2EQ71LA, 2EQ71HA, 1SZ87DA, 1SZ87LA, 1VD87LA, 1VX87LA, 1AK87QQ, 1QM89WJ, 1YS87LA, 1CL89NP, 1RM87LA, 1RW27JA, 1NM87BA, 1BM87LA |
| Germ cell tumours | 04249, 05480, 06111, 06113, 06170, 06173, 06177, 04241, 06177, 04244, 04245, 06122, 06125, 06309, 06390, 06073, 06191, 06261, 06262, 18171, 18172, 18173, | 1OT87LA, 1OT91LA, 1QH87LB, 1QM87LA, 1QM89DA, 1QM89LA, 1RB87DA, 1RB87LA, 1QM91LB, 1QM89WJ, 1RB89RA, 1RD89DA, 1RD89LA, 1RD89RA, 1GX87QF, 1QP87LA, 1QM87WJ, 1GX87LA, 1SF91LA, 1SF89PF, 1NF87LA, 1FA87CA |
| Carcinomas | 05480, 05486, 06023, 06113, 06133, 06134, 06135, 06161, 06168, 06170, 06173, 06174, 06175, 06177, 06185, 06186, 06217, 07157, 07170, 03132, 03159, 03165, 03166, 03167, 03168, 03196, 04537, 04538, 05025, 05170, 05171, 05182, 05183, 05188, 05193, 05480, 06111, 06113, 06162, 06177, 06185, 06186, 07156, 07170, 07171, 07194, 01130, 01131, 01132, 01133, 01134, 01143, 01144, 01199, 01302, 02007, 02083, 02127, 06150, 06151, 06152, 06153, 06168, 06180, 06073, 06191, 06261, 06262, 05304, 05142, 05485, 01205, 02082, 18171, 18172, 18173, 03125 | 1ET87LA, 1OT87LA, 1PE87PF, 1NV89DA, 1YD87LA, 1YU87LA, 1FU59CA, 1FU59HA, 1FA87BA, 1FA87CA, 1FA91LA, 1FU87NZ, 1FU87PZ, 1FU87QT, 1PE87DA, 1PE87LA, 1YA87LA, 1YB87LA, 1YC87LA, 1YD87LA, 1YF87LA, 1YG87LA, 1YR87LA, 1YS87LA, 1YT87LA, 1YV87LA, 1YZ87LA, 1FA91LA, 1FU89NZ, 1FU91NZ, 1GW87LA, 1GW87DA, 1GR87NW, 1GR87DA, 1GJ87LA, 1NM87DA, 1NM87LA, 1NK87BA, 1NK87DA, 1NK87DN, 1NK87LA, 1GJ87LA, 1GJ87QB, 1GX87LB, 1GX87QF, 1NQ87BA, 1NQ87CA, 1NQ87DA, 1NQ87LA, 1NQ87PF, 1NT87LA, 1YM87LA, 1FU87NZ, 1FU87PZ, 1FU87QT, 1YB87LA, 1YC87LA, 1YD87LA, 1YF87LA, 1YG87LA, 1YR87LA, 1YS87LA, 1YT87LA, 1YU87LA, 1YV87LA, 1YW87LA, 1YX87LA, 1YZ87LA, 1GK87LA, 1GK87DA, 1YK87LA, 1YL87LA, 1YM87DA, 1YM87GB, 1YM87LA, 1YM87UT, 1CQ87LA, 1FV87NZ, 1FV87PZ, 1ET87BA, 1FX87BA, 1ET87LA, 1FX87LA, 1GB87BA, 1GB87LA, 1GD87BA, 1GD87LA, 1GE87BA, 1GE87LL, 1GJ87LA, 1GJ87QB, 1NK87BA, 1NK87DA, 1NK87DN, 1NK87DX, 1NK87DP, 1NK87DY, 1NK87LA, 1NK87RE, 1NK87RF, 1NK87TF, 1NK87TG, 1NM87BA, 1NM87DA, 1NM87DE, 1NM87DF, 1NM87DN, 1NM87DX, 1NM87DY, 1NM87LA, 1NM87PN, 1NQ87BA, 1NQ87CA, 1NQ87DA, 1NQ87LA, 1NQ87PF, 1NQ87PN, 1NT87LA, 1GM87DA, 1GM87LA, 1GR87DA, 1GT87DA, 1GT87NW, 1GT87QB, 1GV87DA, 1GV87LA, 1GW87DA, 1GW87LA, 1GX87LB, 1GX87QB, 1GX87QF, 1DA87LA, 1FU89NZ, 1RB89LA, 1FU91NZ, 1PM91LA, 1RM91DA, 1RM91DA, 1FM87LA, 1FM87VW, 1NV89LA, 1YE87LA, 1OJ87LA, 1FB87LA, 1FT91LA, 1RB87DA, 1FM89LA, 1FU89PZ, 1OK87VZ, 2GT71BA, 1FM89VW, 1FN89LA, 1GR87QB, 1GR89QB, 1FC87LA, 1ED87LA, 1VC87LA, 1NQ89LH, 1CP89LA |
| Other | 05486, 06161, 06170, 03196, 05171, 05188, 05193, 05480, 02007, 02083, 02127, 06073, 05142, 01205, 02082, 18171, 18172, 18173 | 1OJ89LA, 1OJ87DA, 1NM87DA, 1NM87LA, 1NK87BA, 1NK87DA, 1NK87DN, 1NK87LA, 1GW87LA, 1GW87DA, 1OJ87VC, 1OT87DA, 1OT87LA, 1NM87BA, 1NM87DA, 1NM87DE, 1NM87DF, 1NM87DN, 1NQ87BA, 1NQ87CA, 1NQ87DA, 1NQ87LA, 1NQ87PF, 1OJ87DA, 1OJ87LA, 1OJ87VC, 1OJ87VK, 1OT87DA, 1OT87LA, 1NA87BA, 1NA87DA, 1NA87DB, 1NA87QB, 1NF87BA, 1NF87DA, 1NF87LA, 1HA87DA, 1HA87LA, 1KT87LA, 1SZ87LA, 1YD87LA, 1GR87DA, 1GR87LA, 1WV87LA, 1QM91LB, 1VX87LA |
| **Radiation-related services** | | |
|  | 08513, 08514, 08515, 08551, 08552, 08554, 08555, 08564, 08512, 08545, 08546, 08562, 08563, 08519, 08518, 08517, 08504, 08510, 08511, 08518, 08520, 08553, 08565, 20158, 20159, 20160, 41031, 08501 | 1AC27JX, 1AE27JX, 1AF27JA, 1AF27JX, 1AG27JX, 1AJ27JX, 1AN27JA, 1AN27JX, 1AP27JX, 1AW27JA, 1AW27JX, 1BA27JX, 1BB27JX, 1BD27JX, 1CZ26JA, 1CZ27JA, 1CZ27JX, 1EA27JA, 1EQ27JA, 1FG27JA, 1FJ27JA, 1FP27JA, 1FU27JA, 1FX27JA, 1GE27JA, 1GJ27JA, 1GT27JA, 1GT27JX, 1HZ27JA, 1MD27JA, 1MF27JA, 1MG27JA, 1MH27JA, 1MJ27JA, 1MZ27JA, 1NA27JA, 1NF27JA, 1NK27JA, 1NM27JA, 1NQ27JA, 1NQ27JX, 1NT27JA, 1OA27JA, 1OA27JX, 1OB27JA, 1OD27JA, 1OE27JA, 1OJ27JA, 1PB27JA, 1PC27JA, 1PC27JX, 1PE27JA, 1PE27JX, 1PG27JA, 1PM27JA, 1PQ27JA, 1QE27JA, 1QG27JA, 1QM27JA, 1QT27JA, 1QT27JX, 1RB27JA, 1RF27JA, 1RM27JA, 1RN27JA, 1RN27JX, 1RS27JA, 1RW27JA, 1RZ27JA, 1SC27JA, 1SH27JA, 1SL27JA, 1SZ27JA, 1TX27JA, 1TZ27JA, 1VX27JA, 1VZ27JA, 1WZ27JA, 1YF27JA, 1YS27JA, 1YT27JA, 1YV27JA, 1YV27JA |

| **Supplementary Table S2.** Univariate and multivariable model estimates of mean length of stay ratios | | | |
| --- | --- | --- | --- |
| Characteristic | | Univariate mean LOS ratio (95% CI) | Multivariable mean LOS ratio (95% CI)^a^ |
| Sex | | | |
|  | M | 1.10 (1.01 to 1.19) | 1.04 (0.97 to 1.12) |
|  | F | Ref. | Ref. |
| Cancer type | | | |
|  | Leukemia | Ref. | Ref. |
|  | Lymphoma | 0.55 (0.49 to 0.61) | 0.57 (0.51 to 0.64) |
|  | CNS | 0.62 (0.56 to 0.69) | 0.67 (0.60 to 0.75) |
|  | Neuroblastoma | 0.67 (0.58 to 0.78) | 0.66 (0.57 to 0.77) |
|  | Retinoblastoma | 0.14 (0.11 to 0.18) | 0.13 (0.11 to 0.18) |
|  | Renal tumours | 0.64 (0.53 to 0.77) | 0.64 (0.53 to 0.77) |
|  | Hepatic tumours | 0.81 (0.61 to 1.08) | 0.82 (0.62 to 1.09) |
|  | Bone tumours | 0.54 (0.46 to 0.62) | 0.57 (0.49 to 0.67) |
|  | Soft tissue sarcomas | 0.39 (0.33 to 0.45) | 0.42 (0.36 to 0.50) |
|  | Germ cell tumours | 0.52 (0.43 to 0.64) | 0.57 (0.46 to 0.70) |
|  | Carcinomas | 0.29 (0.25 to 0.34) | 0.38 (0.32 to 0.44) |
|  | Other | 0.51 (0.26 to 0.98) | 0.76 (0.39 to 1.46) |
| Age group at admission | | | |
|  | <1 | 1.78 (1.58 to 2.01) | 1.75 (1.56 to 1.97) |
|  | 1–4 | Ref. | Ref. |
|  | 5–9 | 0.80 (0.75 to 0.86) | 0.85 (0.79 to 0.91) |
|  | 10–14 | 0.85 (0.78 to 0.92) | 0.96 (0.88 to 1.05) |
|  | 15–19 | 0.80 (0.73 to 0.88) | 1.03 (0.94 to 1.14) |
| COVID-19 period | | | |
|  | Pre-pandemic | Ref. | Ref. |
|  | Post-pandemic | 0.83 (0.79 to 0.88) | 1.01 (0.94 to 1.10) |
| Material deprivation index, quintile | | | |
|  | 1 – least | Ref. | Ref. |
|  | 2 | 1.01 (0.92 to 1.12) | 1.01 (0.92 to 1.11) |
|  | 3 | 1.16 (1.05 to 1.28) | 1.17 (1.06 to 1.29) |
|  | 4 | 1.11 (1.00 to 1.22) | 1.11 (1.01 to 1.23) |
|  | 5 – most | 1.23 (1.11 to 1.37) | 1.24 (1.12 to 1.37) |
| Health region type at admission | | | |
|  | University | Ref. | Ref. |
|  | Peripheral | 0.37 (0.31 to 0.44) | 0.44 (0.37 to 0.53) |
|  | Intermediate, northern, or remote | 0.42 (0.38 to 0.46) | 0.40 (0.37 to 0.44) |
| Day of the week upon admission | | | |
|  | Monday | Ref. | Ref. |
|  | Tuesday | 0.79 (0.75 to 0.83) | 0.78 (0.75 to 0.82) |
|  | Wednesday | 0.96 (0.92 to 1.01) | 0.99 (0.94 to 1.04) |
|  | Thursday | 0.80 (0.76 to 0.84) | 0.80 (0.77 to 0.84) |
|  | Friday | 1.06 (1.00 to 1.12) | 1.06 (1.00 to 1.12) |
|  | Saturday | 1.38 (1.27 to 1.49) | 1.39 (1.28 to 1.50) |
|  | Sunday | 1.36 (1.26 to 1.47) | 1.38 (1.28 to 1.49) |
| Month of admission | | | |
|  | January | Ref. | Ref. |
|  | February | 1.04 (0.96 to 1.12) | 1.04 (0.97 to 1.12) |
|  | March | 0.94 (0.87 to 1.01) | 0.94 (0.87 to 1.01) |
|  | April | 0.94 (0.87 to 1.01) | 0.95 (0.89 to 1.02) |
|  | May | 0.87 (0.81 to 0.94) | 0.88 (0.82 to 0.94) |
|  | June | 0.97 (0.90 to 1.04) | 0.98 (0.91 to 1.05) |
|  | July | 0.88 (0.81 to 0.94) | 0.90 (0.84 to 0.97) |
|  | August | 0.91 (0.84 to 0.98) | 0.93 (0.86 to 1.00) |
|  | September | 0.95 (0.88 to 1.02) | 0.96 (0.89 to 1.03) |
|  | October | 0.98 (0.91 to 1.06) | 0.99 (0.92 to 1.07) |
|  | November | 0.92 (0.85 to 0.99) | 0.94 (0.88 to 1.01) |
|  | December | 0.90 (0.83 to 0.96) | 0.91 (0.85 to 0.98) |
| Calendar time | | | |
|  |  | 0.995 (0.994 to 0.996) | 0.996 (0.995 to 0.998) |
| ^a^ Models adjusted for Sex, cancer type, age group at admission, pandemic period, quintile of material deprivation, health region type at admission, day of week admitted, month of admission, and calendar time. | | | |
